# A dual proteomics analysis of paired cerebrospinal fluid and plasma from patients with neurodegenerative diseases

**DOI:** 10.64898/2026.09.02.26361977

**Authors:** Isabelle Kowal, Sonja W. Scholz, Jacob Epstein, Bryan J. Traynor, Laura E. Danielian, Ying Hao, Jody Crook, Ziyi Li, Katelyn Porter, Brian Sellers, Thomas J. Langowski, Marisa N. Denkinger, Nicholas J. Ashton, Kendall Van Keuren-Jensen, Mark R. Cookson, Justin Y. Kwan, Yue A. Qi, Allison Snyder

## Abstract

**INTRODUCTION:** Understanding concordance across biofluids and platforms is critical for understanding neurodegenerative biomarkers results and translating them into clinical use; yet systematic comparisons remain limited. To address this gap, we performed large-scale proteomic profiling of patient-paired plasma and CSF to characterize cross-modal relationships.

**METHODS:** We profiled paired plasma and CSF from 67 individuals using SomaScan 11K and NULISAseq CNS panels. Disease severity was assessed with the CDR+NACC FTLD-M Global Score.

**RESULTS:** We identified 269 SomaScan and 18 NULISA proteins with significant cross-biofluid correlation. Cross-platform concordance within biofluids was strong. NEFL, NPTX2, TREM2, and CHIT1 demonstrated consistent cross-platform agreement. Associations with disease severity were compartment-specific, with decreased NPTX2 in CSF, increased NEFL and GFAP in plasma, and decreased TREM2 across biofluids.

**DISCUSSION:** Cross-platform consistency supports biomarker robustness, while limited cross-biofluid concordance highlights compartmental biology. Given additional clinical correlations despite varying underlying pathology, these findings may point to shared neurodegenerative disorder pathways.

## 1 BACKGROUND

Neurodegenerative diseases are a major cause of behavioral, cognitive, and motor dysfunction resulting in significant functional impairment in aging populations. Reliable biomarkers are crucial for diagnosis, disease monitoring, and clinical trials.^1^ Autopsy studies have historically been the gold standard, but fail to provide definitive diagnosis in life. Cerebrospinal fluid (CSF) studies have offered a reasonable in vivo measure of CNS pathology but are limited because of perceived invasiveness and limited availability. In the case of Alzheimer’s disease, the development of plasma biomarkers has transformed the diagnostic landscape.^2^ However, plasma biomarkers are once again removed from the primary pathology compartment with a complex list of medical conditions and other factors that impact target levels and interpretation. Furthermore, some biomarkers may be CNS-specific and not reliably detected in plasma. An important step in biomarker discovery lies in understanding the shared and compartment-specific findings and ensuring that they are robust despite varying techniques and assays.

While some biomarkers, such as neurofilament light chain (NEFL), have shown consistent relationships across biofluids^3^, many proteins demonstrate inconsistent trends. Blood–brain barrier function, peripheral protein sources, and clearance mechanisms likely contribute to these inconsistencies. A clearer understanding of cross-biofluid relationships is essential for translating CSF-based discoveries into clinically useful blood biomarkers.

Advances in proteomic technologies have enabled large-scale proteomic profiling, though they rely on fundamentally different binding mechanisms. Platforms such as SomaScan provide broad, high-throughput coverage of the proteome^4^ by utilizing aptamers, which are chemically modified, single-stranded DNA sequences designed to bind to specific protein structures. In contrast, targeted approaches such as NULISA (NUcleic acid Linked Immuno-Sandwich Assay) rely on traditional antibody-based detection, utilizing pairs of highly specific antibodies to target proteins with established or suspected relevance to neurodegenerative disease.^5^ Because aptamers and antibodies differ in binding affinities, cross-reactivity, and matrix interference, findings may not be directly comparable across platforms.

Few studies have systematically evaluated both cross-biofluid and cross-platform protein relationships using a cohort with paired CSF and plasma samples.^6^ Such comparisons are important for identifying biomarkers that are both biologically informative and technically robust. Establishing this consistency is a necessary step in bridging the translational divide before downstream validation and clinical translation.

In this study, we analyzed paired CSF and plasma samples from a mixed neurodegenerative disease cohort using two complementary proteomic platforms, SomaScan 11K and NULISAseq CNS Disease Panel 120. Individual assessment of established and emerging neurodegenerative biomarkers across biofluids was performed.^7,8,9,10,11,12,13^ We aimed to (1) assess cross-biofluid protein correlations within each platform, (2) evaluate cross-platform agreement within each biofluid, and (3) incorporate disease severity scores in an exploratory fashion to begin to understand the clinical relevance of established and candidate neurodegenerative biomarkers.

## 2 METHODS

### 2.1 Participants

Enrolled participants in the National Institutes of Health (NIH) IRB-approved protocol “Investigating Complex Neurodegenerative Disorders Related to Amyotrophic Lateral Sclerosis and Frontotemporal Dementia” (clinicaltrials.gov identifier: NCT03225144) greater than 18 years old with available paired spinal fluid and plasma biofluids were included in the analyses. All participants completed clinical evaluations including history, neurological examination, neuropsychological testing, motor assessment, and biofluid collection. Participants were recruited from May 2021 to June 2024. Diagnoses included frontotemporal dementia-amyotrophic lateral sclerosis (FTD-ALS) spectrum disorders, presymptomatic pathogenic variant carriers, Alzheimer’s disease, and Lewy body diseases. Final diagnoses were determined by consensus review. See supplemental data for more details.

### 2.2 Clinical assessments

Symptom duration was calculated in months based on a review of medical records and participant and informant interviews to determine the earliest estimated time of symptom onset. Disease severity was assessed using a derivation of the Clinical Dementia Rating^®^ plus the National Alzheimer’s Coordinating Center Frontotemporal Lobar Degeneration scales that incorporated a motor domain, CDR +NACC FTLD-M, using the global score.^14^

### 2.3 Biospecimen collection

Matched plasma and CSF samples were collected from each participant. Plasma samples were obtained from the upper limb via phlebotomy under non-fasting conditions. Following collection, samples were centrifuged within 2 hours to remove blood cells, and the plasma supernatant was carefully extracted, transferred into EDTA tubes, and stored at −80°C. CSF was collected via lumbar puncture performed in the upright position at the L3/L4 level using a 22-gauge Sprotte needle. Following collection into polypropylene tubes, samples were stored at −80°C. To preserve sample integrity, freeze-thaw cycles were limited to two or fewer before undergoing proteomic analysis.^15^

### 2.4 Study design

We analyzed paired cerebrospinal fluid (CSF) and plasma samples from the mixed neurodegenerative disease cohort. Aliquots of each biofluid were profiled using two advanced proteomic platforms: the SomaScan 11K assay (SomaScan), an aptamer-based capture platform, and NULISA, an antibody-based targeted panel (Supplementary figure S1A).^16,5^

### 2.5 SomaScan 11K Assay

Proteomic profiling was performed using the SomaScan 11K assay (SomaScan), a multiplex aptamer-based proteomic platform. 75μl aliquots of CSF or plasma were used for the assay. Raw fluorescence intensity data were processed using SomaLogic’s standard data processing pipeline, which includes hybridization-control normalization, median-signal normalization, calibration, and plate normalization to minimize technical variation. Processed data were reported as relative fluorescence units (RFUs), reflecting relative protein abundance. Detailed descriptions of the assay and its normalization procedures have been published previously.^16, 17^

### 2.6 NULISAseq CNS Disease 120 Panel

Targeted protein profiling was conducted using NULISAseq^TM^ CNS Disease Panel 120 (NULISA) on the paired plasma and CSF samples at the Fluid Biomarker Program, Banner Sun Health Research Institute. This antibody-based assay targets 127 proteins associated with neurodegenerative disease processes.^5^ Assays were performed at the Fluid Biomarker Program, Banner Sun Health Research Institute according to manufacturer protocols. Raw sequencing read counts were normalized using intra- and inter-plate controls and log2-transformed to generate NULISA Protein Quantification (NPQ) values prior to data delivery.

### 2.7 Quality control and data filtering

#### SomaScan data

Using preprocessed data from SomaLogic’s standard pipeline for both plasma and CSF datasets, protein RFU values below the buffer control threshold were set to missing (i.e.,NA). Because of varying overall protein abundances, filtering was performed separately for each biofluid. Proteins with missing values in more than one-third of samples (n>22) were excluded, corresponding to a signal-to-noise ratio greater than 1.15. We chose to use a threshold that was more stringent than that used in previously reported studies^18^ to remove the prominent low-signal peak in CSF, which likely reflects background noise. All subsequent analyses were conducted using the filtered proteins common to both biofluids.

#### NULISAseq data

Proteins with less than 50% target detectability (n>33), were excluded. Subsequent analyses were conducted using the filtered proteins common to both biofluids.

### 2.8 Statistical analysis

All statistical analyses were conducted in R version 4.4.1 using RStudio. To account for potential confounding by age and sex, protein abundance values from both platforms were adjusted using linear regression. For each protein, separate linear models were fit for CSF and plasma, with protein abundance as the dependent variable and age and sex as independent variables. Age was modeled as continuous and sex as categorical. Residuals from these models, representing protein abundance independent of age and sex, were extracted and used as adjusted protein values for all downstream analyses. Covariate adjustment was performed separately for CSF and plasma measurements to preserve biofluid-specific effects.

To assess the relationships between plasma and CSF protein levels within and across platforms, Spearman correlation coefficients were calculated for each shared protein using adjusted values. Correlation strength (ρ) and corresponding p-values were obtained on a per-protein basis. To account for multiple comparisons across proteins, p-values were adjusted using the Benjamini-Hochberg false discovery rate (FDR) method. Statistical significance was defined as p_adj_<0.05. Heatmaps were generated from the top predictors identified in the univariate regression analyses using the ordinal R package. For each biofluid-platform dataset, proteins were ranked by the nominal p-value from the ordinal regression model for CDR + NACC FTLD-M Global Score, and the top features were selected for visualization. The ordinal regressions were carried out using the clm R package. Patient columns were ordered by increasing global score, and protein values were displayed after row-wise z-score scaling to enable comparison of relative expression patterns across patients.

## 3 RESULTS

### 3.1 Cohort description

The analysis cohort (n=67) consisted of male (n=30) and female (n=37) individuals ranging in age from 29 to 82 years (median=64, interquartile range [IQR] = 56.5–71). The cohort was predominantly White (88.06%). Refer to supplementary data for additional diagnostic details (Table S1). Refer to table one for a cohort summary (Table 1).

**Table 1.** Patient demographic and clinical data summary.

| **Characteristic** | **N** | **Percentage (%)** |
| --- | --- | --- |
| **Sex** |  |  |
| Male | 30 | 44.78 |
| Female | 37 | 55.22 |
| **Age** |  |  |
| <50 | 11 | 16.42 |
| 50-59 | 10 | 14.93 |
| 60-69 | 27 | 40.29 |
| ≥70 | 19 | 28.36 |
| **Race** |  |  |
| White | 59 | 88.06 |
| African American | 3 | 4.47 |
| Asian | 2 | 2.99 |
| Multiracial | 2 | 2.99 |
| Other | 1 | 1.49 |
| **Disease severity, CDR +NACC FTLD-M Global Score** |  |  |
| 0 | 13 | 19.40 |
| 0.5 | 11 | 16.42 |
| 1 | 33 | 49.25 |
| 2 | 7 | 10.45 |
| 3 | 3 | 4.48 |

### 3.2 Evaluating cross-biofluid protein correlation within proteomic platforms

Inspection of the SomaScan data revealed a lower signal-to-noise ratio in CSF compared to plasma (Supplementary figure 2A–2B). After filtering, 9,657 proteins remained in plasma and 3,702 proteins remained in CSF. For SomaScan, 3698 proteins (38.3%) were shared across biofluids, and the other 5959 proteins (61.7%) were unique to plasma. After filtering the NULISA data, 119 proteins remained for plasma and 105 proteins remained for CSF (Supplementary figure 2C–2D). For NULISA, 102 proteins (83.6%) were shared, 17 (13.9%) were plasma-specific, and 3 (2.5%) were CSF-specific (Figure 1A).

**Figure 1:**
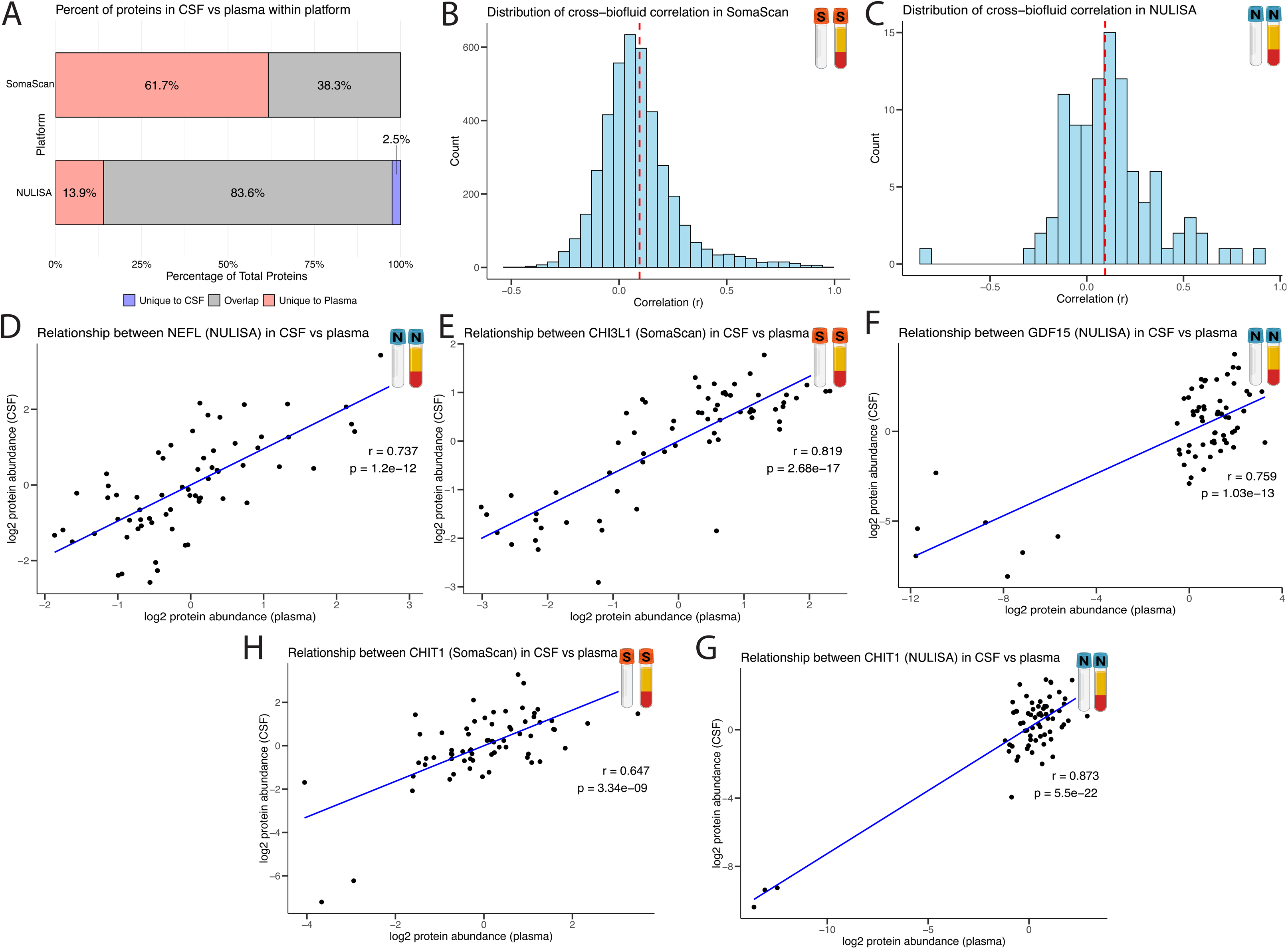
Cross-biofluid correlation analysis within each proteomics platform. (A) In SomaScan, 38.3% of proteins overlapped between CSF and plasma, and in NULISA, 83.6% of proteins overlapped between CSF and plasma. (B) SomaScan CSF and plasma proteins had a median cross-biofluid correlation of 0.0664. (C) NULISA CSF and plasma proteins had a median cross-biofluid correlation of 0.0953. (D) In NULISA, NEFL is positively correlated in CSF and plasma with p_adj_=3.05E-11. (E) In SomaScan, CHI3L1 is positively correlated in CSF and plasma with p_adj_=5.76E-15. (F) In NULISA, GDF15 is positively correlated in CSF and plasma with p_adj_ =3.51E-12. (G) In SomaScan, CHIT1 is positively correlated between CSF and plasma with p_adj_=2.16E-07. (H) In NULISA, CHIT1 is positively correlated between CSF and plasma with p_adj_=5.61E-20.

For SomaScan, 269 proteins showed significant cross-biofluid correlation (p_adj_<0.05). The proportion of significantly correlated proteins was higher for NULISA (17.6% of shared proteins) compared to SomaScan (7.3% of shared proteins).

Cross-biofluid correlations were modest for SomaScan and NULISA, with median ρ values of 0.0664 and 0.0953, respectively (Figure 1B–1C). Neurofilament light chain (NEFL) showed significant positive correlation across biofluids in the targeted NULISA platform (ρ=0.737, p_adj_=3.05E-11; Figure 1D) but no cross-biofluid correlation in SomaScan (ρ=0.033, p_adj_=0.958; Supplementary Figure 3A).In SomaScan, Chitinase-3-like protein 1 (CHI3L1, also known as YKL-40), demonstrated strong cross-biofluid correlation (ρ=0.819, p_adj_=5.76E-15; Figure 1E) but no correlation in NULISA (ρ=0.180, p_adj_=0.368; Supplementary Figure 3B). In NULISA, growth differentiation factor 15 (GDF15) showed significant correlation between CSF and plasma (ρ=0.759, p_adj_=3.51E-12; Figure 1F) but no cross-biofluid correlation in the SomaScan platform (ρ=0.264, p_adj_=0.242; Supplementary Figure 3C). In both SomaScan and NULISA, the glial marker chitinase-1 (CHIT1) was positively correlated between CSF and plasma (ρ=0.647 and 0.873, p_adj_=2.16E-7 and 5.61E-20, respectively; Figure 1G–1H).

### 3.3 Strong cross-platform protein correlation within biofluid compartment

In the plasma, 102 proteins (85.7% of NULISA plasma analytes) were present in both platforms. In CSF, 63 proteins (60% of NULISA CSF analytes) overlapped (Figure 2A). In plasma, 60 proteins demonstrated significant cross-platform correlation with a median ρ of 0.341 (Figure 2B); in the CSF, 47 proteins were correlated across platforms with a median ρ of 0.421 (Figure 2C).

**Figure 2:**
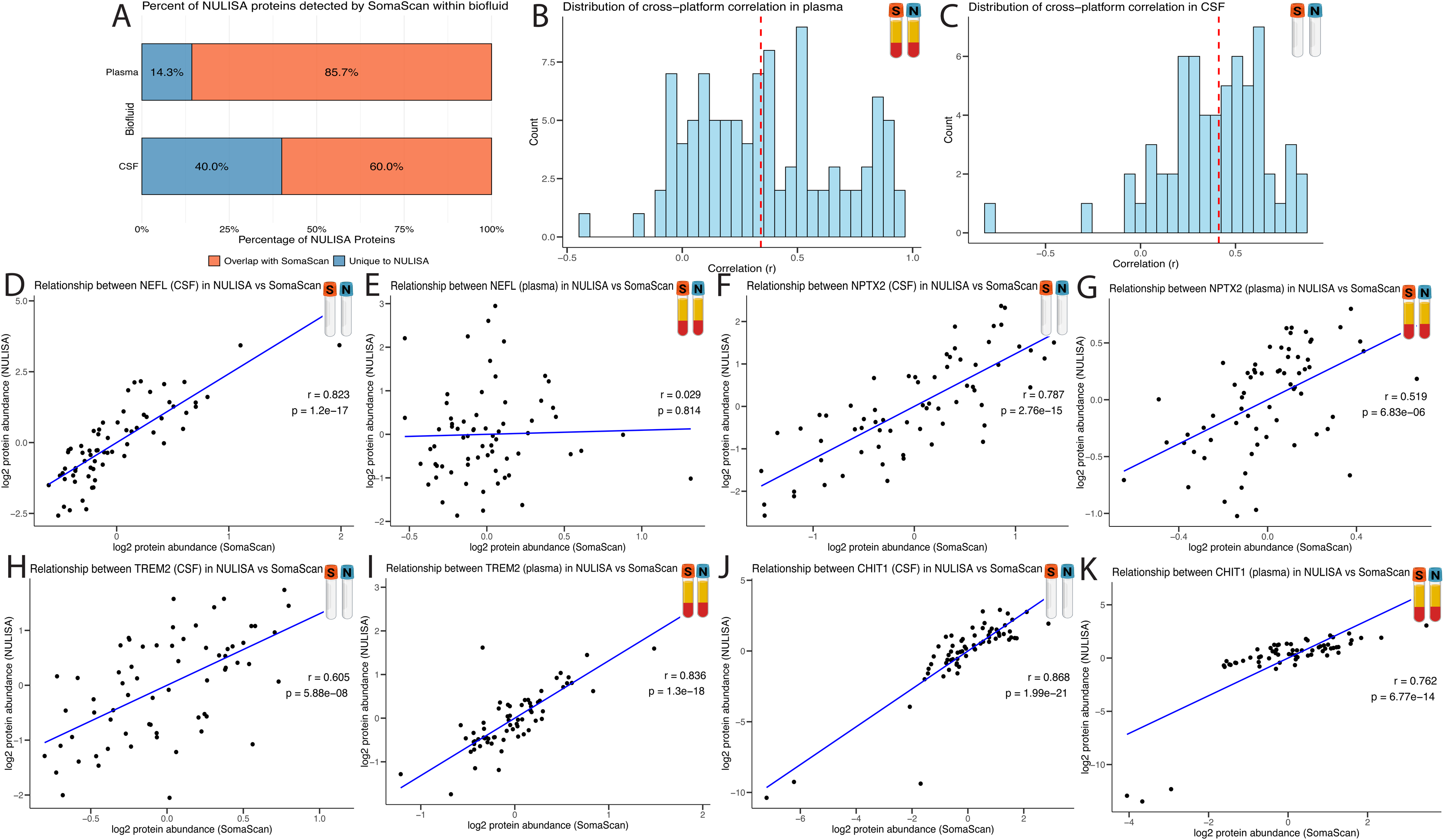
Cross-platform correlation analysis within each biofluid compartment. (A) 85.7% of NULISA plasma proteins are found in SomaScan plasma, and 60% of NULISA CSF proteins are found in SomaScan CSF. (B) Plasma SomaScan and NULISA proteins had a median cross-platform correlation of 0.341. (C) CSF SomaScan and NULISA proteins had a median cross-biofluid correlation of 0.421. (D) CSF NEFL is significantly correlated between platforms with =3.79E-16. (E) Plasma NEFL has no significant correlation between platforms with p_adj_=0.845. (F) CSF NPTX2 is significantly correlated between platforms with p_adj_=3.47E-14. (G) Plasma NPTX2 is significantly correlated between platforms with p_adj_=2.40E-05. (H) CSF TREM2 is significantly correlated between platforms with p_adj_=2.65E-07. (I) Plasma TREM2 is significantly correlated between platforms with p_adj_=1.02E-17. (J) CSF CHIT1 is significantly correlated between platforms with p_adj_=1.25E-19. (K) Plasma CHIT1 is significantly correlated between platforms with p_adj_=4.06E-13

Amongst established and candidate neurodegenerative biomarkers, NEFL showed strong cross-platform correlation in CSF (ρ=0.823, p_adj_=3.79E-16; Figure 2D). However, plasma NEFL had no significant cross-platform correlation (ρ=0.033, p_adj_=0.958; Figure 2E). In contrast, both CSF neuronal pentraxin 2 (NPTX2) (ρ=0.787, p_adj_=3.47E-14; Figure 2F) and plasma NPTX2 (ρ=0.519, p_adj_=2.40E-05; Figure 2G) showed a strong cross-platform correlation. Another consistent candidate biomarker with positive cross-platform correlation in CSF and plasma was triggering receptor expressed on myeloid cells 2 (TREM2) (ρ=0.605 and 0.836, p_adj_=2.65e-07 and 1.02E-17, respectively; Figure 2H–2I). In addition to its strong cross-biofluid correlations described above, plasma and CSF CHIT1 were significantly correlated across SomaScan and NULISA platforms (ρ=0.868 and 0.763 and p_adj_=1.25E-19 and 4.06E-13, respectively; Figure 2J–2K).

### 3.4 Comparison of platform concordance of cross-biofluid correlations

Overall, cross-biofluid correlations were largely concordant across platforms (Figure 3A). The dotted line (y=x) represents equal cross-biofluid correlation strength between SomaScan and NULISA, with deviation from the line indicating platform-specific differences in cross-biofluid comparisons. We highlighted a set of 16 pre-selected proteins that have either been previously established or associated with neurodegenerative diseases (Table 2). Only one, protein C-reactive protein (CRP), had a large platform discordance, defined as a standardized perpendicular distance from the line exceeding 2 standard deviations (|z|>2). Notably, CRP exhibited a strong positive cross-biofluid correlation in SomaScan but a strong negative correlation in NULISA. All candidate biomarkers with significant cross-biofluid correlation within either platform are summarized visually (Figure 3B). The complete table of all 61 proteins measured by both NULISA and SomaScan in CSF and plasma is provided in the supplementary data (Table S2).

**Figure 3:**
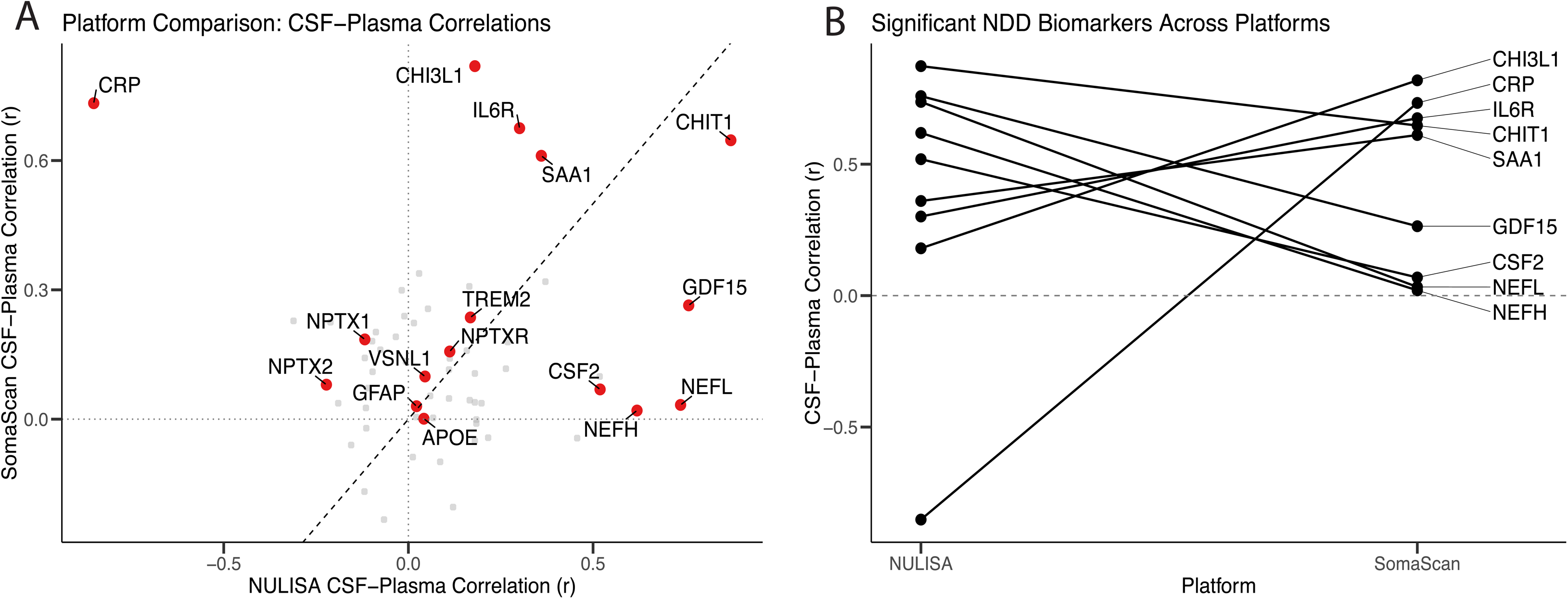
Cross-biofluid correlation platform concordance. (A) Highlighted proteins from Table 2 are represented with red dots, and the dotted line represents perfect concordance. (B) The cross-platform relationship of proteins that had significant correlation in either or both proteomic platforms.

**Table 2.** Selected neurodegenerative-associated proteins panel.

| Gene | NULISaseq CNS Disease Panel<br>120<br>r (p) | SomaScan 11K Assay<br>r (p) |
| --- | --- | --- |
| APOE | 0.042 (0.738) | 0.001 (0.997) |
| CHI3L1 | 0.180 (0.144) | **0.819 (2.68E-17)** |
| CHIT1 | **0.873 (5.50E-22)** | **0.647 (3.34E-09)** |
| CRP | **-0.852 (5.67E-20)** | **0.733 (1.79E-12)** |
| CSF2 | **0.519 (6.90E-06)** | 0.069 (0.578) |
| GDF15 | **0.759 (1.03E-13)** | 0.264 (0.031) |
| GFAP | 0.022 (0.858) | 0.030 (0.812) |
| IL6R | 0.301 (1.30E-02) | **0.675 (3.87E-10)** |
| NEFH | **0.619 (2.35E-08)** | 0.020 (0.872) |
| NEFL | **0.737 (1.20E-12)** | 0.033 (0.790) |
| NPTX1 | -0.118 (0.344) | 0.185 (0.135) |
| NPTX2 | -0.222 (0.071) | 0.080 (0.521) |
| NPTXR | 0.112 (0.336) | 0.157 (0.205) |
| SAA1 | **0.36 (2.80E-03)** | **0.611 (3.94E-08)** |
| TREM2 | 0.168 (0.173) | 0.236 (0.054) |
| VSNL1 | 0.045 (0.719) | 0.099 (0.424) |

### 3.5 Exploring the association of protein abundance with disease severity

We next evaluated associations between protein abundance and disease severity as measured by CDR + NACC FTLD-M global scores across biofluids and platforms (e.g. NULISA CSF, NULISA plasma, SomaScan CSF, and SomaScan plasma). The top five proteins associated with increasing disease severity are summarized with a heatmap (Figure 4A). The selected proteins correspond to the strongest associations observed across the full set of analyzed proteins and are representative of the upper range of effect sizes in each dataset.

**Figure 4:**
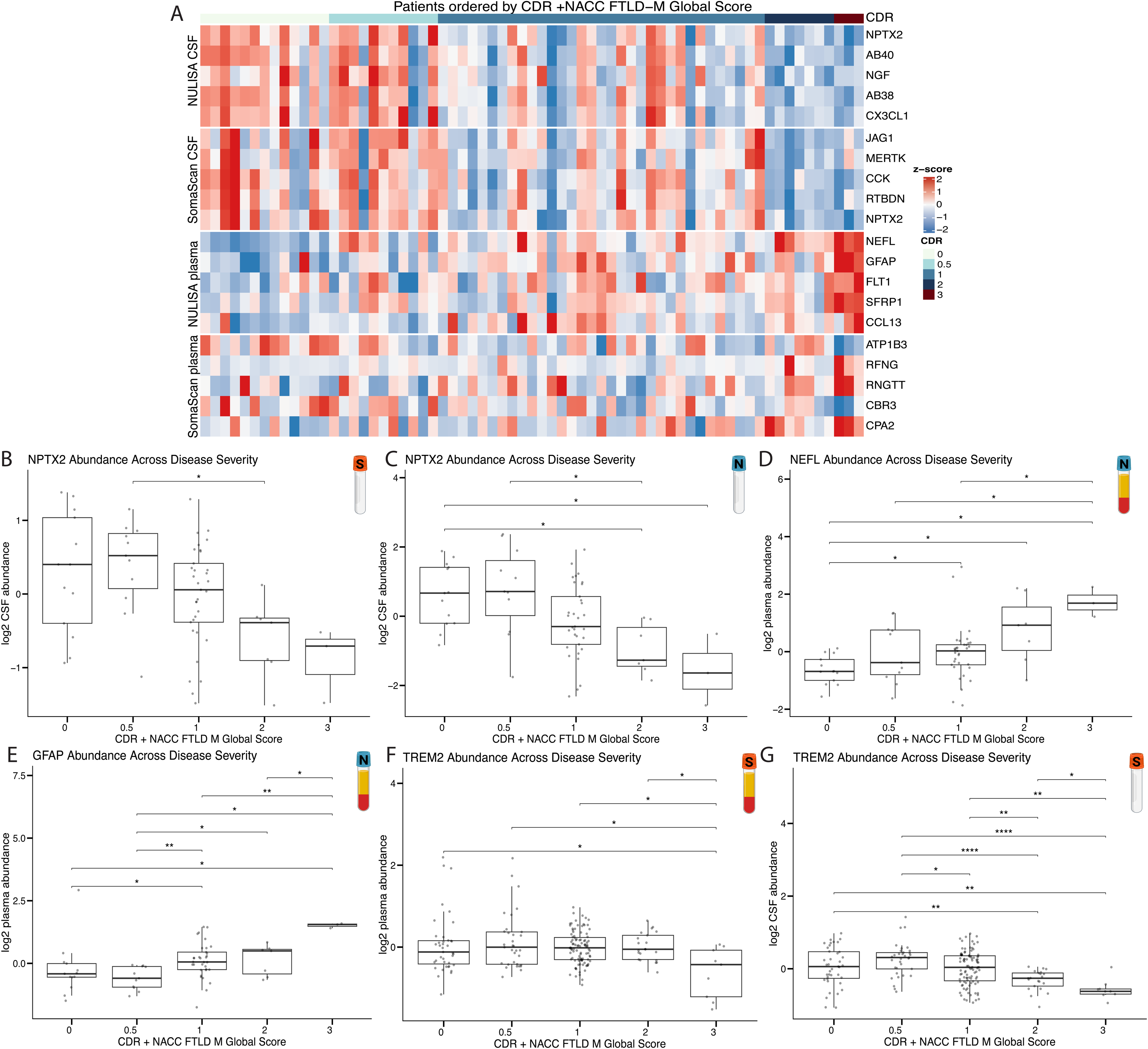
CDR + NACC FTLD M Global Score enables disease severity measure. (A) Heatmap of top 5 proteins in CSF and plasma of both SomaScan and NULISA platforms. Patients (columns) are ordered by increasing CDR FTLD-M global score, and rows are groups by platform and biofluid. (B) CSF NPTX2 levels decrease as disease severity increases in both NULISA and (C) SomaScan platforms. (D) Plasma NEFL and (E) GFAP levels increase as disease severity increases in NULISA. (F) CSF TREM2 and (G) plasma TREM2 levels decrease in SomaScan platform.

Among established neurodegenerative biomarkers, NPTX2 emerged as a top hit in CSF across both platforms. Consistent with this, NPTX2 abundance decreased significantly with increasing CDR FTLD-M global score in both SomaScan and NULISA (Figure 4B–4C). Additional proteins identified in NULISA CSF included Aβ40, Aβ38, CX3CL1, and NGF, which have reported associations with neurodegenerative disease. In the NULISA plasma, NEFL and GFAP showed significant increases in abundance with increasing CDR score. FLT1, SFRP1, and CCL13, which are not established neurodegenerative biomarkers, also demonstrated similar trends and can be potential candidates to explore in future studies (Figure 4D–4E). Finally, TREM2, though not a top 5 predictor of disease severity, demonstrated decreasing protein abundance with increasing disease severity across biofluids in the SomaScan platform (Figures 4F–4G).

## 4 DISCUSSION

Neurodegenerative diseases are biologically complex and even in clear-cut clinical syndromes, multiple underlying pathologies frequently coexist. While biomarkers for Alzheimer’s Disease, and to a lesser extent alpha-synucleinopathies, have advanced considerably, validated tools for the remaining neurodegenerative pathologies remain limited (PMID: 36864661). Establishing reliable, scalable biomarkers is an essential first step toward reframing neurodegenerative disorders as biological diseases, enabling earlier diagnosis and intervention; accordingly, it is critical to evaluate whether current biomarker platforms perform consistently across analytical platforms and biofluid compartments.

In this study, we performed a systematic comparison of cross-biofluid and cross-platform protein relationships in paired CSF and plasma samples using two complementary proteomic platforms. Our findings highlight key axes that govern biomarker translation: biological variability across compartments and technical variability across assays. Overall, we demonstrated a strong agreement across platforms within the same biofluid can coexist with limited concordance between biofluids, implying that analytical reproducibility does not necessarily imply biological equivalence across compartments.

We observed strong cross-platform agreement for overlapping proteins within both CSF and plasma, supporting technical reproducibility of modern proteomic techniques. Several established and candidate biomarkers, including NPTX2, TREM2, and CHIT1^19,20,21^, demonstrated consistent cross-platform correlations, suggesting that these analytes are robust to differences in assay design. This reproducibility provides confidence that independent proteomic approaches can capture similar biological signals and support their continued use in biomarker discovery pipelines.

In contrast, correlations between CSF and plasma protein levels were generally modest. Rather than representing a limitation alone, these findings likely reflect biologically meaningful compartmentalization. This finding is consistent with previous studies suggesting that protein concentrations in the peripheral do not always directly reflect central nervous system processes (PMID: 42054495).^22^ Differences in blood-brain barrier permeability, protein clearance, and peripheral production of proteins may all contribute to the limited correspondence between these compartments. As a result, while some proteins demonstrated measurable cross-biofluid relationships, many CNS-derived signals may remain more readily detectable in CSF than in plasma. These results reinforce the concept that not all CNS-derived signals are readily detectable in plasma and successful translation of biomarkers to blood requires identification of proteins that either cross compartments or reflect systemic responses to CNS pathology.

Despite this discordance, a subset of analytes demonstrated encouraging cross-biofluid relationships. Notably, CHIT1 emerged as a particularly robust candidate, exhibiting strong correlations between CSF and plasma across both platforms. This consistency suggests that CHIT1 may represent a stable marker of neuroinflammatory signaling that is detectable outside the CNS. Similarly, platform-specific cross-biofluid correlations were observed for NEFL, GDF15, and CHI3L1^23,24,25^, further illustrating that certain biological signals may be captured differently depending on assay characteristics. The translation of these glial markers between compartments suggests that certain immune-related signals may function as reliable peripheral markers of neurodegeneration outside the central nervous system and, if centrally derived, may indicate compromise of the blood-brain barrier.^26^

At the same time, variability in widely used biomarkers underscores important translational challenges. NEFL, a widely used marker of neurodegeneration, has been used as outcome measures in clinical trials and longitudinal studies (PMID: 36129998 and PMID: 33827960) and is available clinically for prognostication (PMID: 25934855). Consistent with previous reports, NEFL showed increased CSF and plasma levels with increasing disease severity in the NULISA dataset.^27^ Although NEFL demonstrated strong cross-biofluid correlation in NULISA, no correlation was observed in SomaScan, suggesting that platform-specific differences in assay sensitivity, epitope recognition, or dynamic range for specific analytes may influence cross-biofluid concordance for certain analytes. These findings do not diminish the potential utility of NEFL as a biomarker, but highlight the need for assay standardization and careful interpretation when comparing results across platforms or clinical studies.

The evaluation of cross-platform concordance provides an additional lens through which to assess biomarker readiness for translation. In our study, we found strong agreement across platforms in both CSF and plasma NPTX2. Similarly, TREM2 and CHIT1 showed consistent cross-platform correlations, supporting their potential as reliable markers of microglial activation. TREM2, a microglial receptor involved in innate immune signaling and previously implicated in neurodegenerative disease risk^28^, has previously been reported to decrease in CSF in FTD but not plasma.^12^ Recent studies have linked higher baseline plasma GDF15 levels to steeper global cognitive decline and accelerated neurodegeneration in FTD.^24^ These proteins that demonstrate stability across platforms may represent particularly promising candidates for further development as they are less susceptible to technical and biological variability.

Lastly, on an exploratory basis, we sought to determine the clinical utility of these markers by relating them to a measure of disease severity. NPTX2 is a synaptic protein whose CSF levels have been reported to decrease in neurodegenerative disorders.^29,30^ Consistent with prior literature, we found that CSF NPTX2 decreased with increasing CDR in both assays. The concordance of these findings with previously reported biomarker patterns provides internal validation of our dataset and supports the reliability of the observed proteins in clinical context. In the plasma, NEFL and GFAP increased with worsening disease. In combination, these markers have been explored in composite frameworks (e.g., GFAP/NEFL ratios) to capture distinct aspects of neurodegeneration and improve disease characterization.^31^ These clinical correlations highlight potential disease agnostic markers of clinical severity and may suggest programs that are relevant to disease biology at varying stages, though their performance may depend on assay and compartment.

Together, these results have important implications for clinical translation. Effects to develop blood-based biomarkers of disease have transformed the detection, diagnosis, study, and treatment of Alzheimer’s disease. Our findings suggest that successful translation requires a dual-validation framework, in which candidate biomarkers demonstrate cross-platform reproducibility and biologically meaningful behavior across compartments. Markers that meet these criteria are more likely to be robust in real-world clinical and research settings.

While these observations provide insight into cross-platform reproducibility and potential relationships across biofluids and clinical scores, several limitations should be considered when interpreting the results. First, this clinical cohort was relatively small, which limits statistical power. Lack of autopsy data limits the ability to draw definite conclusions regarding biomarker performance as it relates to specific disease mechanisms. Importantly, our study included presymptomatic pathogenic variant carriers with disease severity scores of 0, which may not be fully reflective of their biological state. Similarly, the cohort did not include healthy control participants, which restricts interpretation of disease-related protein changes in relation to normal aging. This study was cross-sectional, preventing assessment of longitudinal effects or relationships between protein levels and disease progression. As a result, our analyses primarily reflect associations with disease severity within an affected cohort rather than inferring disease versus normal differences. Furthermore, differences in assay mechanisms and lack of quantitative results may contribute to variability in measured protein concentrations and cross-biofluid correlations. Because not all proteins were measured across both platforms, the direct comparison for certain analytes was limited. Both SomaScan and NULISA are affinity-based detection methods that use aptamers and antibodies, respectively. They are influenced by factors such as epitope accessibility, protein isoforms, and post-translational modifications. Although the concordance observed across platforms provides some reassurance regarding measurement reliability, confirmation using orthogonal methods such as mass spectrometry would further strengthen the confidence in the observed results.

Despite these limitations, the use of paired CSF and plasma samples combined with dual-platform profiling represents a key strength of this study. This study design provides insight into the extent to which protein signals are preserved across both biological fluids and analytical platforms, which is an important consideration for biomarker translation from discovery to clinical application. Overall, this systematic comparison showed that although cross-biofluid correlations were overall modest, there was substantial agreement across platforms within individual compartments, suggesting that independent technologies can capture consistent biological signals. Several candidate neurodegeneration biomarkers reproduced known clinical associations, supporting the validity of the dataset, while exploratory signals in proteins related to microglial activation and neuroinflammation highlight promising markers for further investigation. Future studies incorporating larger cohorts, longitudinal sampling, healthy controls, and orthogonal validation will be essential to confirm these findings and establish any utility of cross-biofluid proteomic biomarkers in neurodegenerative disease.

## Data Availability

All data produced in the present study are available upon reasonable request to the authors

## ACKNOWLEDGEMENTS

This research was supported in part by the Intramural Research Program of the National Institutes of Health (NIH). The contributions of the NIH authors are considered Works of the United States Government. The findings and conclusions presented in this paper are those of the authors and do not necessarily reflect the views of the NIH or the U.S. Department of Health and Human Services.

## CONSENT STATEMENT

The National Institutes of Health Institutional Review Board approved this study.

## REFERENCES

1. Blennow, K. A Review of Fluid Biomarkers for Alzheimer’s Disease: Moving from CSF to Blood. Neurol Ther. 2017 Jul 21;6(Suppl 1):15–24. doi:10.1007/s40120-017-0073-9.

2. Ankeny SE, Bacci JR, Decourt B, Sabbagh MN, Mielke MM. Navigating the Landscape of Plasma Biomarkers in Alzheimer’s Disease: Focus on Past, Present, and Future Clinical Applications. Neurol Ther. 2024 Sep 7;13(6):1544–1557. doi:10.1007/s40120-024-00658-x

3. Hofmann A, Häsler LM, Lambert M, Kaeser SA, Gräber-Sultain S, Obermüller U, et al. Comparative neurofilament light chain trajectories in CSF and plasma in autosomal dominant Alzheimer’s disease. Nat Commun. 2024 Nov 18;15:9982. doi:10.1038/s41467-024-52937-8

4. Kirsher DY, Chand S, Phong A, Nguyen B, Szoke BG, Ahadi S. Current landscape of plasma proteomics from technical innovations to biological insights and biomarker discovery. Commun Chem. 2025 Sep 25;8(1):279. doi:10.1038/s42004-025-01665-1.

5. Feng W, Beer JC, Hao Q, Ariyapala IS, Sahajan A, Komarov A, et al. NULISA: a proteomic liquid biopsy platform with attomolar sensitivity and high multiplexing. Nat Commun. 2023 Nov 9;14:7238. doi:10.1038/s41467-023-42834-x.

6. Dayon L, Cominetti O, Wojcik F, Galindo AN, Oikonomidi A, Henry H, et al. Proteomes of Paired Human Cerebrospinal Fluid and Plasma: Relation to Blood-Brain Barrier Permeability in Older Adults. J Proteome Res. 2019 Mar 1;18(3):1162–1174. doi:10.1021/acs.jproteome.8b00809.

7. Verde F, Otto M, Silani V. Neurofilament Light Chain as Biomarker for Amyotrophic Lateral Sclerosis and Frontotemporal Dementia. Front Neurosci. 2021 Jun 21;15:679199. doi: 10.3389/fnins.2021.679199.

8. Connolly K, Lehoux M, O’Rourke R, Assetta B, Erdemir GA, Elias JA, et al. Potential role of chitinase-3-like protein 1 (CHI3L1/YKL-40) in neurodegeneration and Alzheimer’s disease. Alzheimer’s Dement. 2022 Mar 2;19(1):9–24. doi:10.1002/alz.12612.

9. Walker KA, Chen K, Shi L, Yang Y, Fornage M, Zhou L. Proteomics analysis of plasma from middle-aged adults identifies protein markers of dementia risk in later life. Sci. Transl. Med. 2023 Jul 19;15(705). doi:10.1126/scitranslmed.adf5681.

10. Varghese AM, Ghosh M, Bhagat SK, Vijayalakshmi K, Preethish-Kumar V, Vengalil S. Chitotriosidase, a biomarker of amyotrophic lateral sclerosis, accentuates neurodegeneration in spinal motor neurons through neuroinflammation. J Neuroinflammation. 2020 Aug 6;17(1):232. doi:10.1186/s12974-020-01909-y.

11. Van der Ende EL, Xiao M, Xu D, Poos JM, Panman JL, Jiskoot LC, et al. Neuronal pentraxin 2: a synapse-derived CSF biomarker in genetic frontotemporal dementia. J Neurol Neurosurg Psychiatry. 2020 Jun;91(6):612–621. doi:10.1136/jnnp-2019-322493.

12. Kleinberger G, Yamanishi Y, Suárez-Calvet M, Czirr E, Lohmann E, Cuyvers E, et al. TREM2 mutations implicated in neurodegeneration impair cell surface transport and phagocytosis. Sci. Transl. Med. 2014 Jul 6;6(243). doi:10.1126/scitranslmed.3009093.

13. Lai R, Li B, Bishnoi R. P-tau217 as a Reliable Blood-Based Marker of Alzheimer’s Disease. Biomedicines. 2024 Aug 13;12(8):1836. doi:10.3390/biomedicines12081836.

14. Snyder A, Samra K, Wu T, Russell LL, Farren J, Crook J, et al. Integrating a motor domain enhances disease severity scales in an FTD-ALS spectrum cohort. Alzheimer’s Dement. 2025 Oct 28;21(10):e70786. doi:10.1002/alz.70786.

15. Chia R, Moaddel R, Kwan JY, Rasheed, M, Ruffo P, Landeck N, et al. A plasma proteomics-based candidate biomarker panel predictive of amyotrophic lateral sclerosis. Nat Med. 2025 Aug 19;31(10):2440–2450. doi:10.1038/s41591-025-03890-6

16. Gold L, Ayers D, Bertino J, Bock C, Bock A, Brody EN, et al. Aptamer-Based Multiplexed Proteomic Technology for Biomarker Discovery. PLOS One. 2010 Dec 7;5(12):e15004. doi:10.1371/journal.pone.0015004.

17. Candia J, Cheung F, Kotliarov Y, Fantoni G, Sellers B, Griesman T, et al. Assessment of Variability in the SOMAscan Assay. Sci Rep. 2017 Oct 7;7(1):14248. doi:10.1038/s41598-017-14755-5.

18. Dammer EB, Ping L, Duong DM, Modeste ES, Seyfried NT, Lah JJ, et al. Multi-platform proteomic analysis of Alzheimer’s disease cerebrospinal fluid and plasma reveals network biomarkers associated with proteostasis and the matrisome. Alz Res Therapy. 2022 November 17;14:174. doi:10.1186/s13195-022-01113-5.

19. Hruska-Plochan M, Wiersma VI, Betz KM, Mallona I, Ronchi S, Maniecka Z, et al. A model of human neural networks reveals NPTX2 pathology in ALS and FTLD. Nature. 2024 Feb 14; 626:1073–1083. doi:10.1038/s41586-024-07042-7.

20. Kleinberger G, Yamanishi Y, Suárez-Calvet M, Czirr E, Lohmann E, Cuyvers E, et al. TREM2 mutations implicated in neurodegeneration impair cell surface transport and phagocytosis. Sci. Transl. Med. 2014 Jul 6;6(243). doi:10.1126/scitranslmed.3009093.

21. Vu L, An J, Kovalik T, Gendron T, Petrucelli L, Bowser R. Cross-sectional and longitudinal measures of chitinase proteins in amyotrophic lateral sclerosis and expression of CHI3L1 in activated astrocytes. J Neurol Neurosurg Psychiatry. 2020 Jan 14;91(4):350–358. doi:10.1136/jnnp-2019-321916.

22. Alcolea D, Beeri MS, Rojas JC, Gardner RC, Lleó A. Blood Biomarkers in Neurodegenerative Diseases. Neurology. 2023 Jul 5;101(4):172–180. doi:10.1212/WNL.0000000000207193.

23. Verde F, Otto M, Silani V. Neurofilament Light Chain as Biomarker for Amyotrophic Lateral Sclerosis and Frontotemporal Dementia. Front Neurosci. 2021 Jun 21;15:679199. doi: 10.3389/fnins.2021.679199.

24. Chen C, Paolillo EW, Saloner R, VandeBunte AM, Cadwallader CJ, Lee SY, et al. GDF15 as a Prognostic Biomarker of Cognitive Decline in Frontotemporal Dementia. Alzheimers Dement. 2025 Jan 9;20(Suppl 2):e089335. doi:10.1002/alz.089335.

25. Connolly K, Lehoux M, O’Rourke R, Assetta B, Erdemir GA, Elias JA, et al. Potential role of chitinase-3-like protein 1 (CHI3L1/YKL-40) in neurodegeneration and Alzheimer’s disease. Alzheimer’s Dement. 2022 Mar 2;19(1):9–24. doi:10.1002/alz.12612.

26. Vu L, An J, Kovalik T, Gendron T, Petrucelli L, Bowser R. Cross-sectional and longitudinal measures of chitinase proteins in amyotrophic lateral sclerosis and expression of CHI3L1 in activated astrocytes. J Neurol Neurosurg Psychiatry. 2020 Jan 14;91(4):350–358. doi:10.1136/jnnp-2019-321916.

27. Gendron TF, Heckman MG, White LJ, Veire AM, Pedraza O, Burch AR, et al. Comprehensive cross-sectional and longitudinal analyses of plasma neurofilament light across FTD spectrum disorders. Cell Rep Med. 2022 April 19;3(4):100607. doi:10.1016/j.xcrm.2022.100607.

28. Li L, Zheng X, Ma H, Zhu M, Li X, Sun X, et al. TREM2 in Neurodegenerative Diseases: Mechanisms and Therapeutic Potential. Cells. 2025 Sep 5;14(17):1387. doi:10.3390/cells14171387.

29. Oh HS, Urey DY, Karlsson L, Zhu Z, Shen Y, Farinas A, et al. A cerebrospinal fluid synaptic protein biomarker for prediction of cognitive resilience versus decline in Alzheimer’s disease. Nat. Med. 2025 Mar 31;31:1592–1603. doi:10.1038/s41591-025-03565-2.

30. Hruska-Plochan M, Wiersma VI, Betz KM, Mallona I, Ronchi S, Maniecka Z, et al. A model of human neural networks reveals NPTX2 pathology in ALS and FTLD. Nature. 2024 Feb 14; 626:1073–1083. doi:10.1038/s41586-024-07042-7.

31. Cousins KAQ, Shaw LM, Chen-Plotkin A, Lee EB, Trojanowski JQ, Van Deerlin VM, et al. Plasma GFAP:NfL discriminates FTLD-tau from FTLD-TDP. Alzheimers Dement. 2022 Dec 20;18(Suppl 5):e064010. doi:10.1002/alz.064010.

