## Supplementary Figure 1 for "A dual proteomics analysis of paired cerebrospinal fluid and plasma from patients with neurodegenerative diseases"

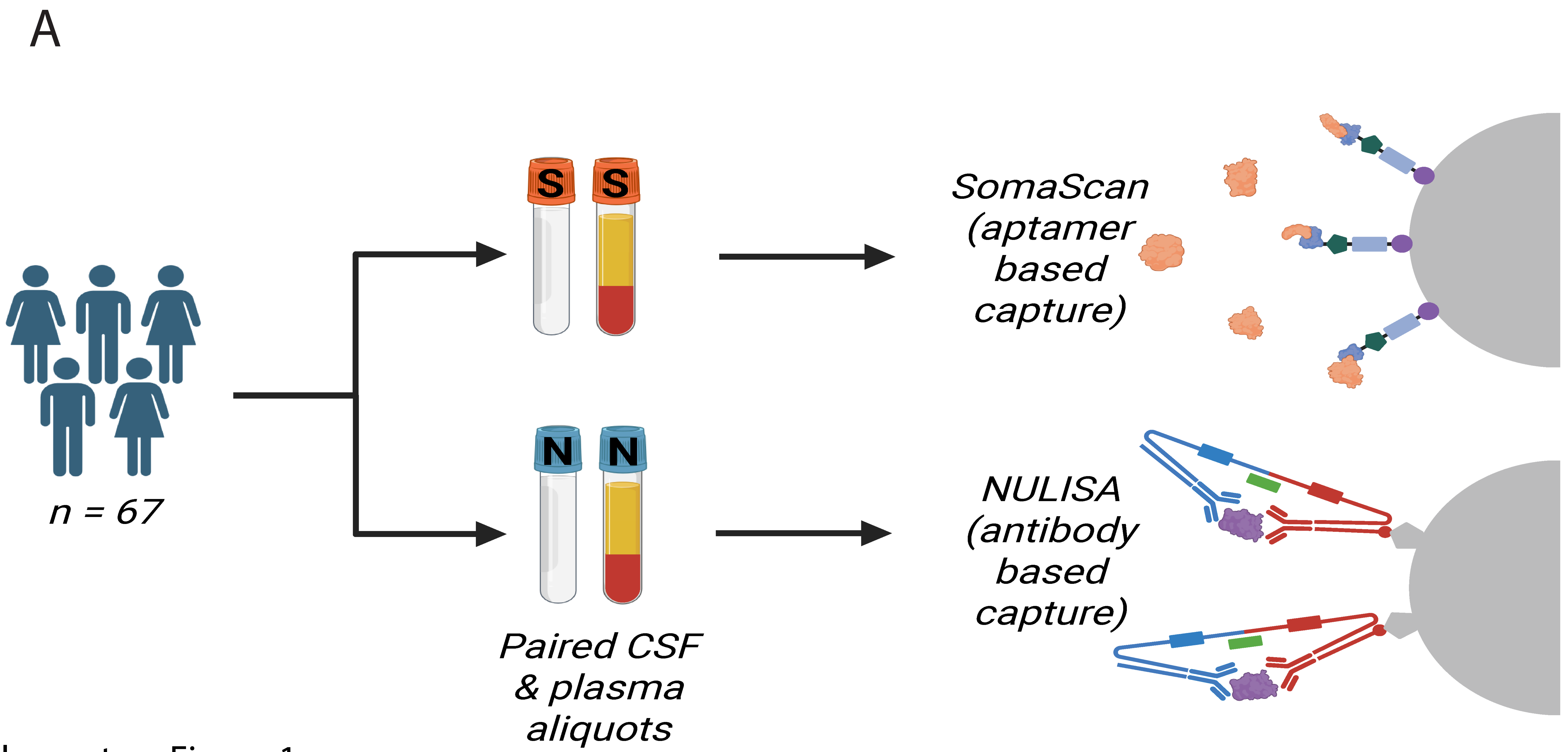


Supplementary Figure 1:
Experiment workflow. CSF and plasma aliquots from 67 patients were sent for SomaScan and NULISA assays.
