## Supplementary Figure 2 for "A dual proteomics analysis of paired cerebrospinal fluid and plasma from patients with neurodegenerative diseases"

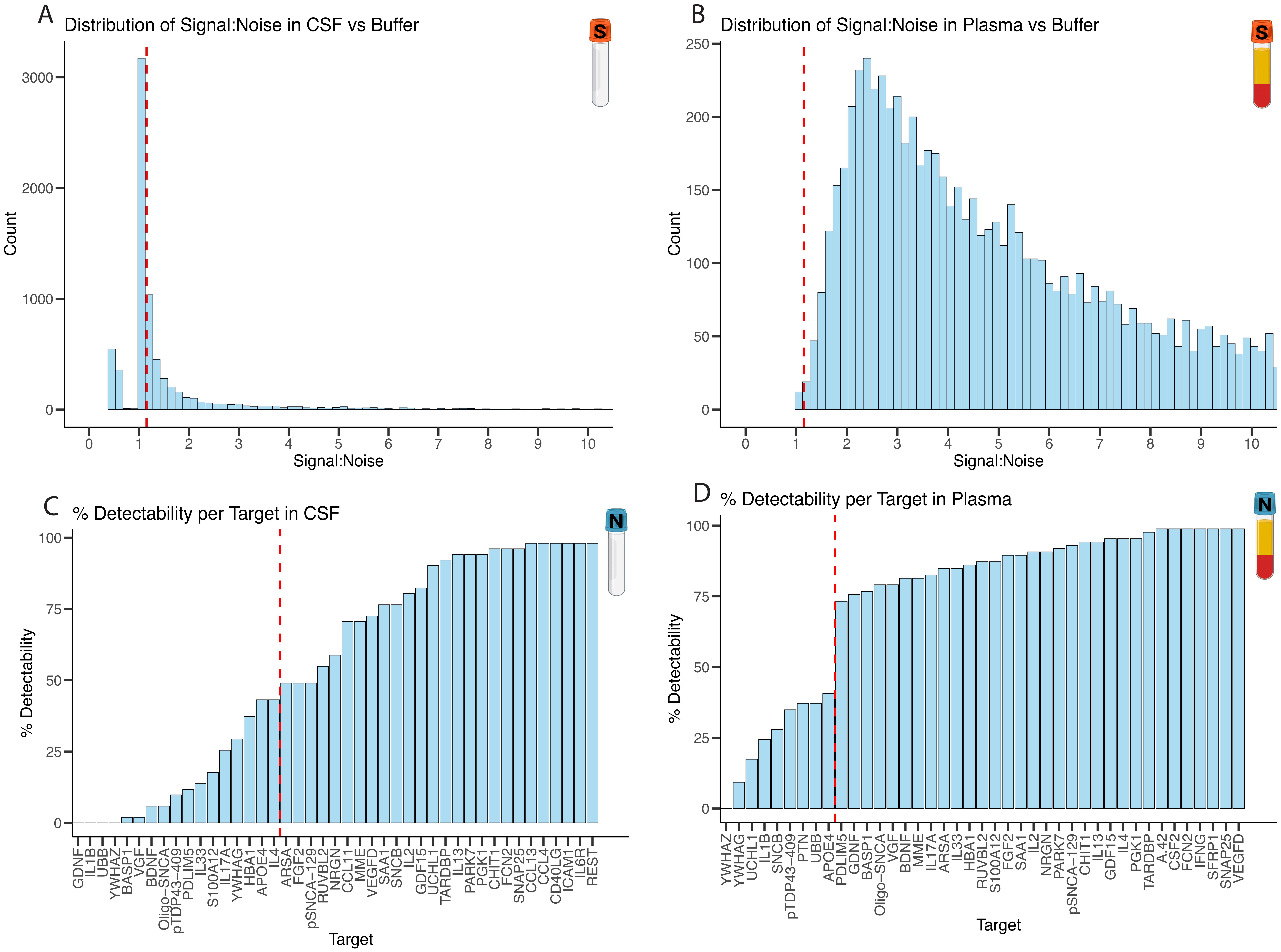


Supplementary Figure 2:
QC and data filtering. (A) Signal to noise ratio of CSF to buffer in SomaScan is very low. (B) Signal to noise ratio of plasma to buffer in SomaScan is very high. A cut off of 1.15 was applied to all SomaScan data. (C) Targets with less than 100% detectability are depicted. CSF proteins below 50% detectability were removed. (D) Targets with less than 100% detectability are depicted. Plasma proteins below 50%
