## Supplementary Figure 3 for "A dual proteomics analysis of paired cerebrospinal fluid and plasma from patients with neurodegenerative diseases"

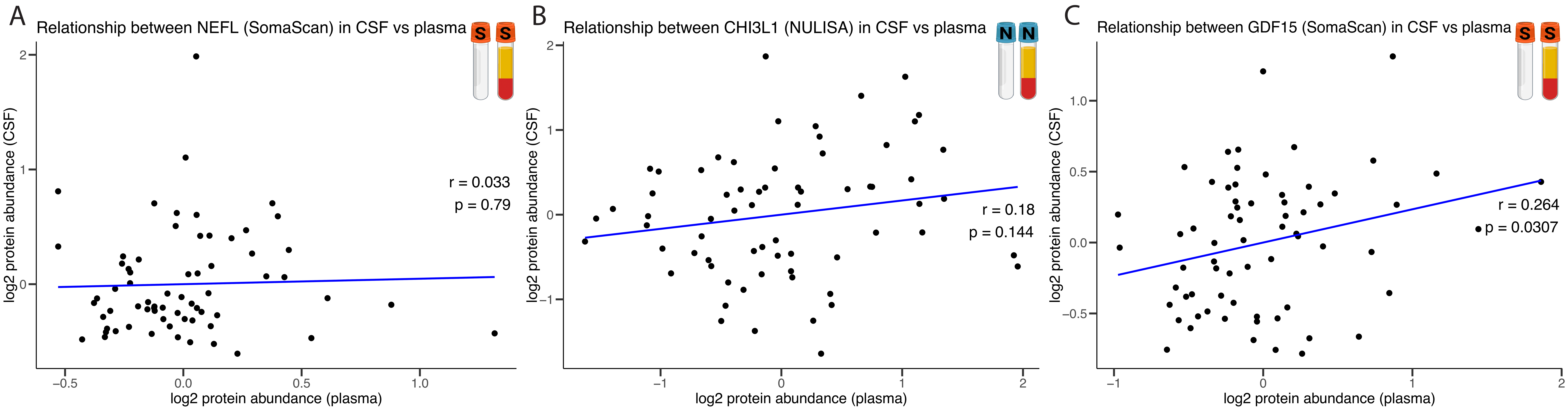


Supplementary Figure 3:
Cross-biofluid correlation analysis within each proteomics platform. (A) In SomaScan, NEFL shows no correlation between CSF and plasma with padj=0.958. (B) In NULISA, CHI3L1 shows no correlation between CSF and plasma with padj=0.368. (C) In Somascan, GDF15 shows no correlation between CSF and plasma with padj=0.242.
