## Supplementary Table 1 for "A dual proteomics analysis of paired cerebrospinal fluid and plasma from patients with neurodegenerative diseases"

**Supplementary Table 1. Complete patient diagnosis summary**

| Diagnosis | N | Percentage (%) |
| --- | --- | --- |
| Frontotemporal dementia  Amyotrophic lateral sclerosis (ALS)  Presymptomatic  Alzheimer's disease (AD)  Alpha-synucleinopathies (PD, DLB, MSA)  Mild cognitive impairment (MCI)  Mild behavioral impairment (MBI) | 9  25  11  11  5  4  2 | 13.43  37.31  16.42  16.42  7.46  5.97  2.99 |
