## Supplementary Table 2 for "A dual proteomics analysis of paired cerebrospinal fluid and plasma from patients with neurodegenerative diseases"

**Supplementary Table 2. Complete cross-biofluid protein data**

| **Gene** | **NULISAseq CNS Disease Panel 120** | | **SomaScan 11K Assay** | |
| --- | --- | --- | --- | --- |
|  | r(p) | p_adjust | r(p) | p_adjust |
| ACHE | 0.165 (0.182) | 0.422 | 0.308 (0.011) | 0.124 |
| ANXA5 | 0.198 (0.108) | 0.335 | 0.037 (0.763) | 0.952 |
| APOE | 0.042 (0.738) | 0.827 | 0.001 (0.997) | 0.999 |
| BACE1 | -0.066 (0.598) | 0.753 | -0.233 (0.058) | 0.354 |
| CALB2 | 0.012 (0.92) | 0.939 | -0.088 (0.48) | 0.866 |
| CCL2 | 0.180 (0.144) | 0.368 | 0.039 (0.757) | 0.952 |
| CCL22 | 0.371(2.00E-03) | 0.015 | 0.319 (0.008) | 0.099 |
| CD63 | -0.076 (0.539) | 0.697 | 0.161 (0.193) | 0.659 |
| CHI3L1 | 0.180 (0.144) | 0.368 | 0.819 (2.68E-17) | 5.69E-15 |
| CHIT1 | 0.873 (5.50E-22) | 5.61E-20 | 0.647 (3.34E-09) | 2.14E-07 |
| CNTN2 | 0.216 (0.079) | 0.269 | -0.043 (0.728) | 0.94 |
| CRH | -0.018 (0.883) | 0.919 | 0.299 (0.014) | 0.144 |
| CRP | -0.852 (5.67E-20) | 2.89E-18 | 0.733 (1.79E-12) | 1.72E-10 |
| CSF2 | 0.519 (6.90E-06) | 8.13E-05 | 0.069 (0.578) | 0.902 |
| CST3 | -0.118 (0.343) | 0.616 | 0.142 (0.250) | 0.722 |
| CXCL1 | 0.112 (0.367) | 0.616 | 0.141 (0.257) | 0.726 |
| CXCL8 | 0.021(0.869) | 0.914 | 0.005 (0.968) | 0.996 |
| ENO2 | -0.311 (0.010) | 0.053 | 0.228 (0.064) | 0.373 |
| FABP3 | -0.011 (0.931) | 0.941 | 0.239 (0.052) | 0.335 |
| FCN2 | 0.184 (0.136) | 0.368 | -0.01 (0.935) | 0.99 |
| FLT1 | -0.155 (0.209) | 0.464 | -0.06 (0.627) | 0.915 |
| FOLR1 | -0.034 (0.784) | 0.86 | 0.191 (0.121) | 0.532 |
| GDF15 | 0.759 (1.03E-13) | 3.51E-12 | 0.264 (0.031) | 0.242 |
| GDI1 | -0.097 (0.436) | 0.645 | 0.11 (0.374) | 0.817 |
| GFAP | 0.022 (0.858) | 0.911 | 0.030 (0.812 | 0.964 |
| GOT1 | 0.053 (0.670) | 0.814 | 0.256 (0.036) | 0.27 |
| ICAM1 | 0.029 (0.816) | 0.885 | 0.338 (5.00E-03) | 0.066 |
| IGF1R | -0.115 (0.354) | 0.616 | 0.026 (0.832) | 0.968 |
| IGFBP7 | 0.110 (0.376) | 0.616 | 0.048 (0.698) | 0.938 |
| IL10 | 0.166 (0.179) | 0.422 | 0.044 (0.721) | 0.94 |
| IL13 | 0.457 (1.01E-04) | 1.03E-03 | -0.044 (0.721) | 0.94 |
| IL16 | 0.067 (0.592) | 0.753 | 0.003 (0.982) | 0.999 |
| IL2 | 0.086 (0.489) | 0.665 | -0.099 (0.424) | 0.834 |
| IL6R | 0.301 (0.013) | 0.062 | 0.675 (3.87E-10) | 2.74E-08 |
| IL9 | 0.180 (0.144) | 0.368 | 0.106 (0.392) | 0.818 |
| KDR | 0.518 (7.17E-06) | 8.13E-05 | 0.099 (0.425) | 0.834 |
| KLK6 | -0.211 (0.087) | 0.287 | 0.225 (0.068) | 0.382 |
| MDH1 | -0.007 (0.956) | 0.956 | 0.134 (0.28) | 0.748 |
| NEFH | 0.619 (2.35E-08) | 4.80E-07 | 0.020 (0.872) | 0.978 |
| NEFL | 0.737 (1.20E-12) | 3.05E-11 | 0.033 (0.790) | 0.958 |
| NPTX1 | -0.118 (0.344) | 0.616 | 0.185 (0.135) | 0.561 |
| NPTX2 | -0.222 (0.071) | 0.263 | 0.080 (0.521) | 0.878 |
| NPTXR | 0.112 (0.336) | 0.616 | 0.157 (0.205) | 0.67 |
| NPY | -0.109 (0.381) | 0.616 | 0.072 (0.562) | 0.897 |
| PARK7 | -0.190 (0.123) | 0.368 | 0.037 (0.768) | 0.954 |
| PGF | 0.264 (0.031) | 0.132 | 0.117 (0.345) | 0.8 |
| PGK1 | -0.098 (0.432) | 0.645 | 0.181 (0.143) | 0.57 |
| POSTN | -0.088 (0.48) | 0.665 | 0.202 (0.101) | 0.479 |
| PRDX6 | 0.158 (0.201) | 0.455 | 0.159 (0.200) | 0.668 |
| SAA1 | 0.36 (2.80E-03) | 0.018 | 0.611 (3.94E-08) | 2.01E-06 |
| SFRP1 | 0.180 (0.144) | 0.368 | -0.05 (0.69) | 0.934 |
| SFTPD | 0.015 (0.904) | 0.932 | 0.223 (0.07) | 0.391 |
| SLIT2 | 0.059 (0.637) | 0.792 | 0.054 (0.662) | 0.926 |
| SMOC1 | 0.121 (0.329) | 0.616 | -0.204 (0.098) | 0.469 |
| TAFA5 | -0.114 (0.36) | 0.616 | -0.021 (0.867) | 0.978 |
| TIMP3 | 0.269 (0.028) | 0.122 | 0.179 (0.147) | 0.577 |
| TREM2 | 0.168 (0.173) | 0.421 | 0.236 (0.054) | 0.344 |
| VCAM1 | -0.119 (0.337) | 0.616 | -0.168 (0.175) | 0.636 |
| VEGFA | 0.110 (0.373) | 0.616 | 0.115 (0.352) | 0.801 |
| VEGFD | 0.184 (0.136) | 0.368 | -0.001 (0.992) | 0.999 |
| VSNL1 | 0.045 (0.719) | 0.827 | 0.099 (0.424) | 0.834 |
